# Efficacy of test and treat with doxycycline on palpable nodules, microfilarial load and *Wolbachia* density in onchocerciasis infected persons in communities with persistent transmission in South-West Cameroon

**DOI:** 10.64898/2026.06.09.26355259

**Authors:** Armelle Forrer, Elizabeth Dibando Obie, Raphael A. Abong, Relindis Ekanya, Abdel J. Njouendou, Theobald Mue Nji, Andrew Amuam, Esum Mathias Eyong, Bertrand L. Ndzeshang, Desmond Akumtoh Nkimbeng, Fanny Fri Fombad, Samuel Teghen, Anicetus Suireng, Ernestine Ebot Ashu, Louise Hamill, Peter Enyong, Joseph D. Turner, Samuel Wanji, Mark J. Taylor

## Abstract

**Introduction:** Onchocerciasis is targeted for elimination with community-directed treatment with ivermectin (CDTI). Alternative strategies are needed in areas where transmission persists despite long-term CDTI and/or are co-endemic with loiasis. This study assessed the efficacy of 35-day treatment with 100mg doxycycline on *Wolbachia* density at 6 months and microfilaridermia and palpable nodules at 30 months post-treatment.

**Methods:** A treatment follow-up study was conducted in 20 high-transmission onchocerciasis communities in a co-endemic loiasis area of South-West Cameroon. Community-based directly observed treatment with 100mg doxycycline was administered to community members aged ≥9 years. *Wolbachia* clearance at 6-months and treatment efficacy on microfilaridermia and palpable nodules were assessed at 30-months post treatment. Factors associated with reductions in microfilaridermia post treatment, including adherence to doxycycline treatment were assessed with mixed-effects logistic regression.

**Results:** Over 92% (2835/3080) of eligible participants took 35 days of 100mg doxycycline over 5 or 6 weeks. This regimen achieved a 62.8% microfilaridermia reduction and 99% palpable nodule reduction in the 720 participants included at follow-up. *Wolbachia* depletion was observed in 92% of skin samples at 6 months post treatment. The most important factor associated with microfilaridermia after 30 months was having missed at least 7 doxycycline consecutive doses (OR 3.11, 95%CI: 1.17-8.26). Incomplete treatment to a lesser extent was not associated with reduced efficacy at follow-up.

**Conclusion:** This large-scale community intervention shows that a 5-week treatment with 100mg doxycycline is feasible and has high curative efficacy against adult *O. volvulus* as measured by the dramatic reduction in the proportion of palpable nodules at 30-months post treatment. The high efficacy shows the tremendous potential of anti-*Wolbachia* drugs as part of the arsenal for onchocerciasis elimination and paves the way for the next generation of anti-*Wolbachia* drugs with shorter treatment courses, which will facilitate the implementation of alternative strategies to accelerate onchocerciasis elimination.

**Key questions box:** *What is already known?:* The mutualistic symbiosis of *Wolbachia* bacteria with their filarial nematode hosts has been exploited as a drug target, to deliver safe curative therapy, which provides an important alternative strategy to facilitate elimination of onchocerciasis transmission. Alternative strategies are required in areas where existing treatment strategies are failing or compromised by safety risks where co-endemic loiaisis occurs.

*What are the new findings?:* Community-based test and treat with doxycycline for 5 weeks achieved a 63% reduction in microfilaridermia and a 99% reduction in palpable adult parasite nodules 30-months post treatment.

*What do those findings imply?:* This study provides the first evidence of a complete curative reabsorption of subcutaneous adult parasite nodules and validates anti-*Wolbachia* therapy as an effective cure for onchocerciasis and a feasible alternative strategy to facilitate onchocerciasis elimination.

## Introduction

Onchocerciasis, also known as river blindness, is a Neglected Tropical Disease (NTD) targeted for elimination in 12 countries by 2030 ^1^. Onchocerciasis is caused by the filarial parasite *Onchocerca volvulus* which is transmitted to humans by blackflies of the genus *Simulium.* It causes visual impairment and irreversible blindness, but most of its burden is due to skin disease manifesting as stigmatising skin conditions and severe itching interfering with sleep and routine activities ^2–4^.

In 2017, there were an estimated 20 million onchocerciasis cases globally, and 240 million people were requiring MDA for onchocerciasis in 2020, over 99% of whom lived in Africa ^5, 6^. Based on morbidity only, the burden of onchocerciasis was estimated at 1.23 million Disability-adjusted Life Years (DALYs), in 2019 ^7^. However, this could be an underestimate as this does not include the impact on mental health and there is increasing evidence that onchocerciasis causes potentially fatal epilepsy in children ^8–11^.

The current WHO-recommended strategy to achieve elimination relies on high coverage (80%) Mass Drug Administration (MDA) with the microfilaricide, ivermectin, over the parasite’s life span (15-17 years), which prevents transmission to blackflies and eventually interrupts transmission ^1, 12^.

However, elimination efforts are hampered by the risk of serious adverse events in individuals harbouring high microfilaraemic *Loa loa* co-infections, and suboptimal coverage of CDTI due to programmatic challenges and negative perception of CDTI ^13–17^. Although CDTI has been an effective control tool and has achieved elimination in some settings, high transmission persists in some areas despite almost two decades of CDTI, including in Cameroon ^13, 16–23^. Alternative strategies are needed to achieve onchocerciasis elimination in areas of high transmission, low acceptability of CDTI as well as in areas co-endemic with loiasis where ivermectin-based treatment cannot be safely deployed due to serious adverse reactions in cases of heavy *L. loa* infection ^15, 21, 24–28^.

The development of a drug killing the adult parasites (a macrofilaricide) has been identified as a critical action to achieve onchocerciasis elimination targets as set out in the WHO NTD road map 2021-2030 ^1^. Test-and-Treat with doxycycline (TTd) is one of the WHO endorsed alternative strategies to ivermectin treatment. Doxycycline is a macrofilaricide targeting the parasite endosymbiont *Wolbachia,* resulting in the permanent sterilisation of adult *O. volvulus* worms and interruption of microfilariae (mf) production, as well as the reduction of the adult lifespan leading to macrofilaricidal activity from 21 months post-treatment ^29–31^. Doxycycline therapeutic performance is superior to other anti-filarial drugs and is completely safe in individuals co-infected with *L. loa*, from which *Wolbachia* is absent ^32–34^. A recent phase II randomized controlled trial to test doxycycline for treatment of onchocerciasis-associated epilepsy decreased acute seizure-related hospitalisation and deaths related to ‘nodding syndrome’ ^35^.

The aim of this study was to assess the efficacy of doxycycline among treated patients during a large-scale community-based implementation of TTd on, *Wolbachia* and microfilarial clearance at 6-months and microfilaridermia reduction and macrofilaricidal activity at 30-months post treatment in high-transmission communities of South-West Cameroon.

## Methods

### Study design

The TTd study was a community-based study consisting of a baseline survey and a follow-up of onchocerciasis infected patients at 6- and 30-months post-treatment. Details about the study design and procedures are available elsewhere ^36, 37^.

In brief, the study was conducted in areas of ongoing onchocerciasis transmission with participants from 20 communities located in the Meme River Basin ^17, 37^. The study consisted of 3 cluster intervention arms: a control arm with 10 communities in the Ndian river basin receiving standard CDTI activities, and two TTd intervention arms with 10 communities each, all located in the Meme River Basin ^17, 37^. Communities in the first intervention arm received TTd, and those on the second intervention arm received TTd combined with vector control consisting in ground larviciding with temephos (Abate, BASF) ^36–38^. Control communities were selected from a different river basin with similar transmission intensity to avoid spillover effects, and intervention communities were allocated to each arm to allow maximum spatial segregation between communities receiving vector control and those which did not ^36^. Communities were engaged and the populations sensitized, censused and screened to identify study participants ^36^. Participants in this study included those who were present in the screening survey at baseline, subsets of infected persons who took the treatment and gave samples for post treatment examination of microfilariae positivity/level, nodule positivity or *Wolbachia* depletion, which were used to assess the efficacy of doxycycline.

### Deviation from the original protocol for follow-up assessments

Due to civil unrest and internal displacement of the study population throughout 2018 and 2019 in the study area, planned follow-up assessments could not be conducted, resulting in major deviations from the original protocol. Security issues prevented conducting entomological monitoring, surveying control communities and follow-up parasitological survey in intervention communities 18-months post treatment. However, follow-up parasitological assessments could be conducted at 6- and 30-months post-treatment, in subsets of participants. Results from this assessment are presented here.

### Testing for onchocerciasis and nodule assessment

*O. volvulus* infection was diagnosed using two 1-mg skin snips taken from each iliac crest with a sterile 2 mm corneo-scleral punch (CT 016 Everhards 2218-15 C, Germany). Mf counts were expressed as mf per skin snip. Participants with at least 1 mf found in any skin snip were identified as positive. Presence and number of palpable nodules were assessed by district hospital nursing staff specifically trained by an expert dermatologist ^36, 39^. Assessment of skin mf and palpable nodules were as follows:

### Assessment of microfilaridermia

For microfilaridermia assessment, participant’s iliac crest was thoroughly cleaned with 70% alcohol and allowed to air dry. Two bloodless skin biopsies (approximately 1 mg each) were taken from the left and right iliac crest using a sterile 2 mm corneo-scleral punch (CT 016 Everhards 2218-15 C, Germany). The snipped areas were dusted with Baneocin antiseptic powder after skin collection.

The skin samples from each participant were placed in two separate wells of a 96 well microtiter plate, containing 100 µl of normal saline (0.9% NaCl solution). The corneo-scleral punch was then sterilised for 5 minutes, transferred into 70% alcohol bath for another 5 minutes and then rinsed in distilled water before re-use. The plates were sealed with parafilm to prevent any spill over or evaporation and skin snip samples incubated at room temperature for 24 hours.

Following the 24 hours incubation, the medium from each sample-well was placed on a clean glass slide and examined under a light microscope at X10 magnification. Emerged mf from each skin snip were counted and reported per skin snip (mf/ss). The skin biopsies and medium were then transferred into 1.5 ml Eppendorf tubes (Eppendorf AG, Hamburg, Germany) containing 80% ethanol (GAPUMA UK Limited) and stored at −20 °C or -80°C for subsequent use.

### Nodule assessment

Trained nurses examined all participants in an enclosed and well-illuminated room. Before the physical assessment for nodules, the characteristic nature of the nodule was described to the participants and each participant was asked if they were aware of any nodules present in their skin. Following their consent, participants were partially undressed and examined using Rapid Epidemiological Assessment (REA) guidelines^17^ with emphasis being laid on the bony prominences of the iliac crest, torso, knees, arms, head and upper trochanter and femur. Onchocercal nodules were identified clinically as firm, non-tender mobile masses beneath the skin. The number of nodules found, and their positions were recorded on anatomical diagrams on the participant recruitment form.

### Test & Treat with doxycycline

The TTd activities consisted of (*i*) diagnosing community members for onchocerciasis with skin snipping and (*ii*) enrolling eligible onchocerciasis-positive participants into treatment and follow-up (eligible to participate: inclusion criteria - resident for ≥5 years, provided informed consent with *O. volvulus* infection ≥ 1mf/skin snip; exclusion criteria – age <9 years, history of chronic disease, *L. loa* microfilaraemia >8000 mf/ml and not pregnant as assessed by a urine pregnancy test strip)^37^.

Participants were treated daily for 5 weeks with 100 mg doxycycline obtained from a Good Manufacture Practice (GMP) Certified pharmaceutical supplier (Chanelle Medical, Loughrea, Co. Galway, Ireland). Treatment was delivered together with a small meal, under Directly Observed Therapy (DOT), i.e. the medication was swallowed in the presence of a community drug distributor (CDD) under direct supervision by the research team or the health system ^39^. Exceptions to DOT were made in case of unavoidable absence and participants were provided capsules and asked to return empty blisters. Treatment uptake, including any deviation from DOT, was recorded daily. At the end of the 35-day treatment course, a 7-day catch-up additional course was offered to participants who missed some treatment days.

Adverse events (AEs) were actively recorded by CDDs supervised by study monitors or the chief of health centre. Participants who experienced mild AEs were advised to continue treatment. Participants were referred to the nearest health centre for assessment and treatment in case of persistence or aggravation of any AE.

### Monitoring of doxycycline treatment efficacy at 6 months post treatment

*Wolbachia* depletion was assessed at 6-months post-treatment as a surrogate biomarker of long-term treatment efficacy, in a subset of participants randomly selected using a random number function in MS Excel among those diagnosed with ≥ 10 mf/mg skin pre-treatment, and who completed the 5-week doxycycline course and reported for examination at 6-months post treatment. The numbers of mf and quantity of *Wolbachia* per mf were determined by microscopy and quantitative polymerase chain reaction (qPCR) targeting *O. volvulus* actin (OvActin; GenBank:M84916.1), and the *Wolbachia ftsZ* gene (*w*Ov*ftsZ*; GenBank: AJ276501), respectively, from skin biopsies ^40^.

### Microfilaridermia, palpable nodule and questionnaire assessments 30 months post-treatment

Following a survey localising the study participants, and a risk assessment of security during the civil unrest period, the follow-up assessment was conducted in Kumba health centre in February and March 2020, 30 months after treatment. Despite remaining insecurity, this time and location were deemed to be safe enough to conduct assessments.

Participants were asked to come to Kumba health centre to answer a questionnaire (basic demographics, displacement, ivermectin uptake since doxycycline treatment, perception of doxycycline, as well as doxycycline vs. ivermectin preference and reasons why, and undergo a skin snip and nodule palpation.

### Data management

Parasitological and questionnaire data were collected with EpiInfo version 3.5.2 (EpiData Association; Odense, Denmark). Further data management and analysis were performed in STATA version 15.0 (StataCorp LP; College Station, United States of America).

Distributions and frequencies of participant characteristics were explored and compared at baseline and follow-up using graphs and boxplots. Pearson χ^2^ was used to test differences in proportions. T-test or Wilcoxon signed rank test were used to compare differences in means across groups, as appropriate.

Self-reported adherence to CDTI at baseline was categorized into: (i) never participated, (ii) participation in up to 75% of rounds and participation into ≥ 75% of rounds while other baseline variables were unchanged ^37^. Observed adherence to doxycycline treatment was assessed based on daily observed treatment uptake and the number of missed doses. The number of consecutive doses missed were calculated and their association with skin infection at follow-up was explored using histograms and summary statistics, after which adherence to doxycycline was categorized into (i) 35 days completed directly, (ii) 35-day treatment course completed with catch-up week (iii) incomplete 35-day treatment course and missed < 7 consecutive doses (iv) incomplete 35-day treatment course and missed ≥ 7 consecutive doses.

For the *Wolbachia* clearance assessments, samples were classified based on the quantitative PCR as follows: (i) *O. volvulus* negative (no signal), (ii) *O. volvulus* positive and *Wolbachia* negative, and (iii) *O. volvulus* positive and *Wolbachia* positive.

### Data analysis

Estimates of microfilaridermia, proportion with nodules and their confidence intervals (CIs) at 30-months follow-up were estimated accounting for the study cluster design and loss to follow-up. To adjust for non-response, the sample was post-stratified on age and sex prior to estimating the proportion of *O. volvulus* microfilaridermia and palpable nodules, assuming the follow-up participants were sampled from the baseline study participants, and a finite population of 3,080 (number of enrolled participants at baseline).

Differences in characteristics between participants present or absent at follow-up as well as differences in proportions or means between independent groups were tested with the χ^2^ test, t-test, Mann Whitney test, or the Kruskal Wallis test, as appropriate. Differences in proportions and means between baseline and 6-months or 30-months follow-up were assessed with the McNemar χ^2^ test, or the Wilcoxon signed rank test. The association between baseline and follow-up mf load was estimated using univariate linear regression with robust estimation.

*Wolbachia* depletion assessments were conducted in all participants included in the 6-months follow-up (primary analysis) and in participants where *O. volvulus* DNA could be detected both at baseline and 6 months was also conducted (secondary analysis).

The effect of adherence to TTd on infection status at follow-up was assessed using mixed-effect logistic regression models, where missing follow-up questionnaire data was addressed using multiple imputation. Data was explored for missingness and missingness mechanism, i.e., missing completely at random (MCAR) or missing at random (MAR) were assessed by regressing missing variables over infection intensity at baseline, a series of socio-demographic and treatment history predictors, and village of residence. A Multiple Imputation (MI) model including outcome at follow-up, all variables associated with infection status at baseline and identified auxiliary variables was developed using MI by Chained Equations (MICE) using augmented logistic regression for binary variables and predictive mean matching for positive continuous variables.

MI using imputation for data with monotone missing pattern was also run. The Fraction of Missing Information (FMI) was checked, and the number of imputations was set to over twice the FMI value. Frequencies and distribution of imputed categorical and continuous variables, respectively, were checked against observed variables. The imputation approach yielding the lowest FMI was selected. Complete case analysis was also run as sensitivity analysis.

The relationship between *O. volvulus* skin infection 30-months post treatment and adherence to the 35-day doxycycline treatment course was assessed using a mixed-effects multivariate logistic regression model. This was built based on expert knowledge, included all variables and interactions associated with infection status at baseline, and adjusted for imbalanced participants characteristics at follow-up. Factors deemed important in affecting infection status at follow-up (baseline mf load, history of ivermectin uptake since the TTd intervention, displacement and adherence to doxycycline treatment) were also included. Several functional forms and categorizations were derived from the original questionnaire variables and used in alternative regression models for sensitivity analysis. The interaction between participation in the catch-up week and the number of consecutive missed doses was checked. Multivariate models including alternate variable sets and functional forms to assess the role of adherence in infection at 30-months post treatment were run. Infection status 30-months post treatment was predicted and plotted using the STATA ‘margins’ and ‘marginsplot’ commands.

### Ethics statement

All participants were explained the study procedures before and after *O. volvulus* infection diagnosis at baseline, before the follow-up assessment and provided informed assent with parental consent (age <18 years) and consent (age ≥ 18 years) before enrollment at each time point. The study was approved by the Liverpool School of Tropical Medicine Research Ethics Committee (reference no. 16-027), the Cameroonian National Ethics Committee for Research on Human Health (approval no. 2016/11/838/CE/CNERSH/SP), and the Division of Health Operations Research within the Cameroonian Ministry of Public Health (approval no. 631-03.17).

## Results

### Characteristics and representativeness of follow-up participants

Due to displacement and insecurity preventing community-based assessments at follow-up, 76.6% (2360/3080) of study participants were absent at follow-up. Yet, all but one (destroyed during the conflict) intervention communities (19/20) were represented at follow-up. Characteristics from all treated participants at baseline (n = 3080) and those who were present at follow-up (n = 720) are presented in Table 1.

**Table 1.**
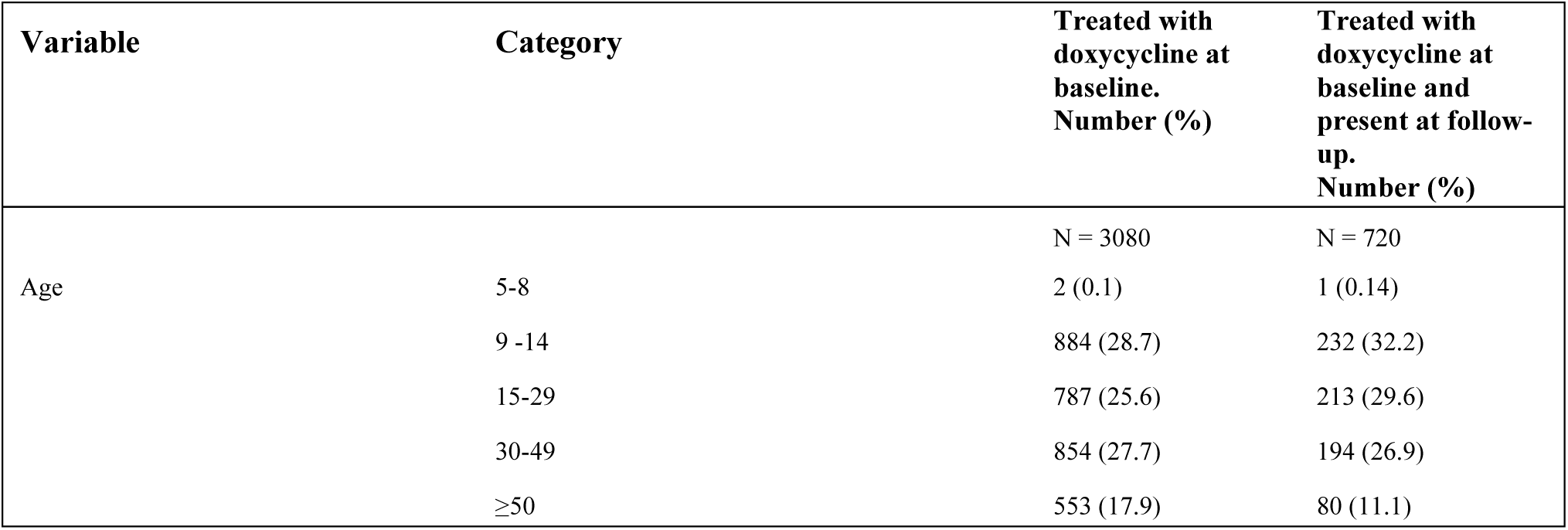
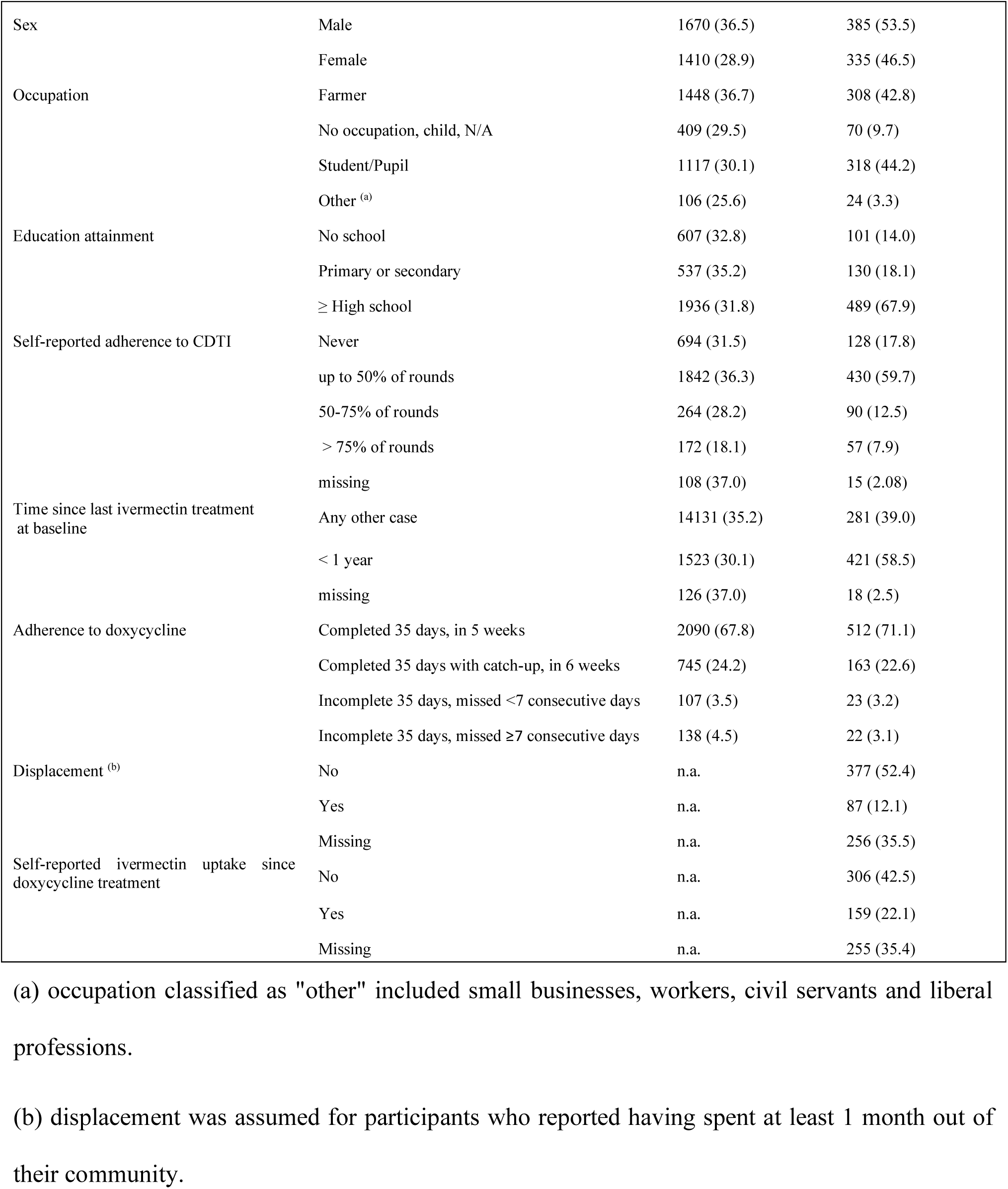
Characteristics of all participants enrolled in doxycycline treatment and those present at 30-months post-treatment follow-up.

**Table 2.**
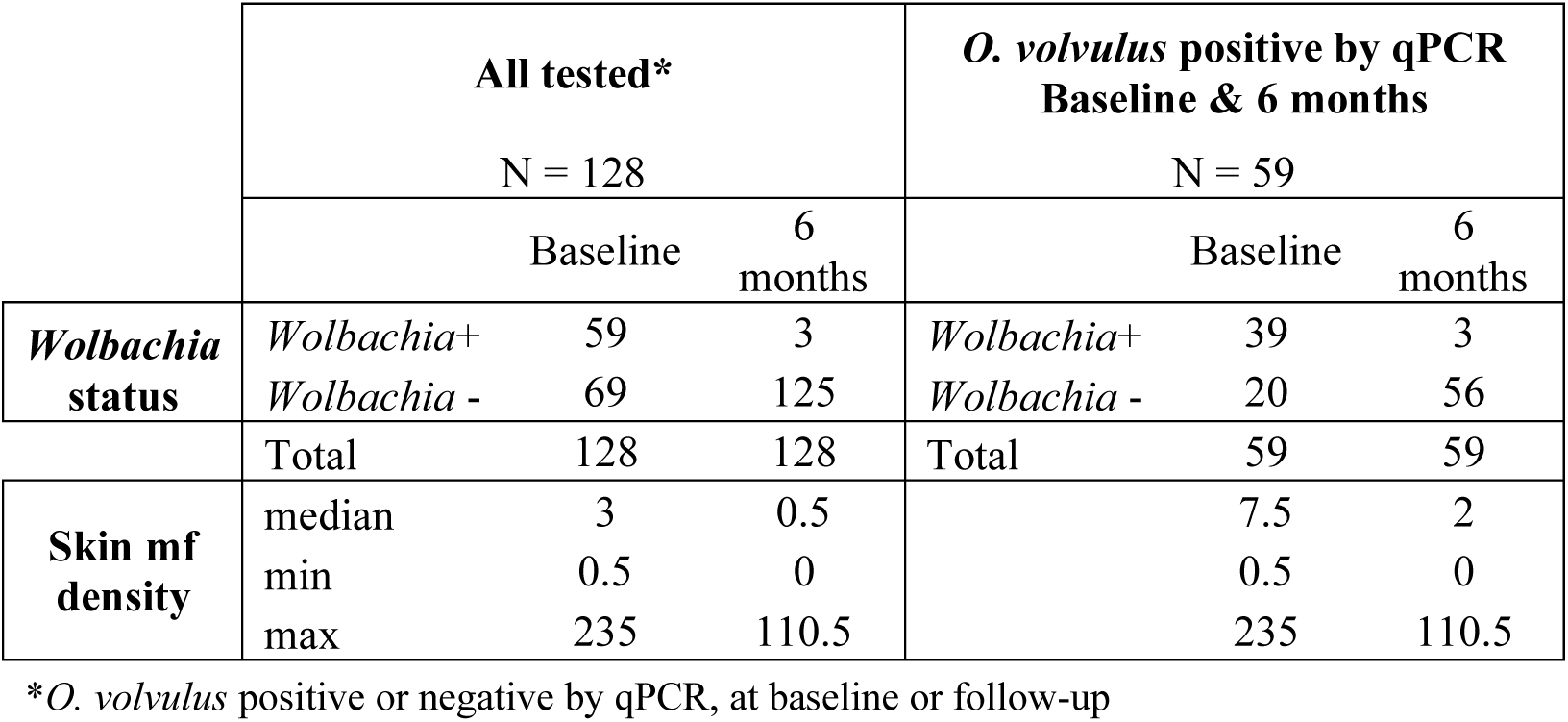
*Wolbachia* status and skin mf density at baseline and follow-up, in all participants tested (n = 128) *vs.* participants tested positive for *O. volvulus* by qPCR both at baseline and 6 months (n = 59).

### Comparing the characteristics of follow-up and baseline participants

Comparison of participants present *vs.* absent at follow-up indicated a similar proportion of men and women (χ^2^ = 0.212, p=0.645). There were significant differences in age group frequencies (χ^2^ = 34.9, p<0.0001), occupation (χ^2^ = 5.926, p=0.015), self-reported adherence to CDTI (χ^2^ = 35.896, p<0.0001), and community (χ^2^ =318.374, p<0.0001).

The DOX-only intervention arm was slightly less represented at follow-up than at baseline (53.5% at follow-up vs. 56.8% at baseline), but microfilarial load at baseline were similar in each arm, i.e., 14.4 mf/g (95%CI: 10.9 – 17.8) in the DOX only arm and 14.1 mf/g (95%CI: 11.0 – 17.2) in the DOX + VC arm (p=0.722).

Mean intensity of onchocerciasis skin infection at baseline among all treated participants (n=3080) was 15.4 mf/g (95%CI: 14.2 – 16.6), and was slightly, but not significantly lower in participants included in follow-up (14.2 mf/g, 95%CI: 11.9 – 16.6) compared to those who were not (15.7 mf/g, 95%CI: 14.3 – 17.1) (Figure 1D) (p = 0.309).

**Figure 1.**
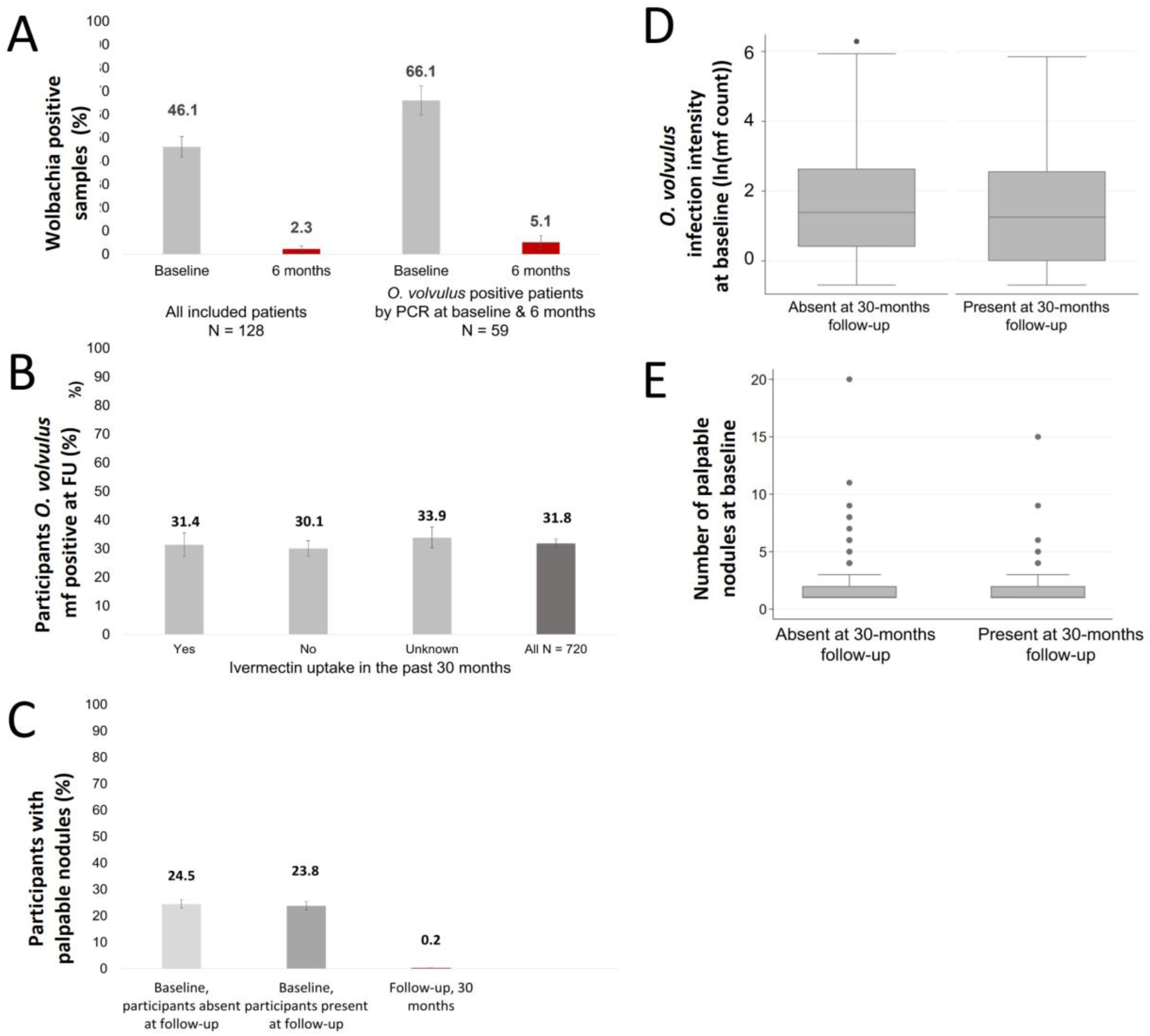
Parasitological and *Wolbachia* assessments. A) Proportion of *Wolbachia* positive snips 6-months post doxycycline treatment in all included participants (N = 128) and participants who were *O. volvulus* actin-positive both at baseline and 6-months follow-up (N = 59). B) Proportion of *O. volvulus* microfilaridermia 30 months post doxycycline treatment, stratified by self-reported ivermectin uptake since doxycycline treatment (light grey bars) and overall (dark grey bar) (N = 720). C) Proportion of participants with palpable nodules at baseline (stratified by absence (N = 2360) *vs*. presence and nodule assessment results (N=719) at 30-month follow-up. D) *O. volvulus* microfilaridermia in participants absent (N = 2360) from, or present at (N = 720), 30-months follow-up. E) Number of palpable nodules in participants at baseline stratified by absence (N = 2360) *vs*. presence and nodule assessment results (N=719) at 30-month follow-up.

Adherence to doxycycline was overall similar between participants present or absent at follow-up, except for those who missed ≥ 7 doses in a row who were significantly less represented at follow-up (OR 0.58, 95%CI 0.37-0.93).

### Follow-up participant characteristics

Only 64.6% (465/720) of participants present at follow-up could be administered the questionnaire due to time and safety constraints. Information on ivermectin uptake since doxycycline treatment and displacement was missing for 95 children aged <12 years and 160 participants aged ≥13 years (355/720, 25.4%). Displacement for a month or more between study surveys was reported by 82/443 (18.5%) aged > 12 years, while about one in eight lived in other locations for 6 months or more. The reported number of months of displacement was similar in the two arms, with means of 2.17 months (range: 0 – 32 months) and 2.58 months (range: 0 – 36) in the TTd only and TTd plus vector control, respectively.

### Adherence to the 35-day doxycycline treatment protocol

Following the community survey, 3,080 eligible participants consented for doxycycline treatment and follow-up. Adherence to the 35-day doxycycline treatment regimen was 92.1% (2,835/3,080) over 5 or 6 weeks. Those rates were slightly higher in the DOC+VC arm (93.3%) than the DOX only arm (91.1%) (chi2=5.09, p=0.024). Of the 3080 participants who completed the treatment course, 2,090 (73.5%) completed it in 5-weeks directly and an additional 745 participants (26.3%) completed the treatment course thanks to the catch-up week. Among participants who missed some treatment days, 49.8% (493/990) missed less than 2 doses, 29.0% and 21.2% (210/990) missed 7 doses or more.

Most participants who missed less than 7 doses (712/780, 91.3%) completed their 35-day treatment week during the catch-up (6th) week, whereas most of those who missed 7 doses or more did not complete their treatment course (177/210, 84.3%). Importantly, among participants who missed at least one dose, about a third (663/990, 67.0%) missed treatment over a maximum number of 2 consecutive days and only 16.3% of participants missed treatment for 7 or more consecutive days.

### Reduction in *O. volvulus* microfilariae and *Wolbachia* from microfilariae at 6-months

*Wolbachia* clearance assessment at 6-months was conducted among 128 patients. At six months post-TTd, 56% (71/128) of the participants had at least one *O. volvulus* mf identified per two skin biopsies (median = 0.5 mf/mg, range 0-110.5 mf/mg) and 73 of 128 samples (57%) were *Ov-act* positive. Skin mf load decreased from 13.10 (95%CI: 7.53-18.58) mf/mg at baseline to 4.43 (95%CI: 2.02-6.85) mf/mg 6 months post doxycycline treatment (p<0.0001).

Of those 73 *O. volvulus* DNA positive samples, 3 were positive for *wOv-ftsz* with *Wolbachia* positivity declining to 4% (3/73), yielding a relative reduction of 95% in *Wolbachia* infection 6 months post doxycycline treatment, as determined by molecular detection.

When assessing *Wolbachia* depletion in the subset of the 59 samples that were *O. volvulus* positive both at baseline and follow-up, the numbers of *Wolbachia* positive samples were 39 and 3 at baseline, and 6 months post doxycycline treatment, respectively, resulting in a relative reduction of 92% in *Wolbachia* infection (McNemar χ^2^ = 34.1, p<0.0001).

### Reduction in microfilaridermia and palpable nodules 30-months post doxycycline treatment

The proportion of microfilaridermia 30-months post-treatment in the 720 study participants present at follow-up was 31.8% (95%CI: 28.8 – 34.8), yielding a relative reduction of 68.2% (all participants were positive at baseline).

In the 720 participants present at follow-up, the mean microfilarial load decreased from 14.4 mf/g (95%CI: 11.9-16.6) at baseline to 3.32 mf/g (95%CI: 2.36 – 4.28) and exhibited a crude positive association with baseline mf load (regression coefficient: 0.07, 95%CI: 0 .02 – 12.6, p = 0.004). In participants who declared no ivermectin uptake since doxycycline treatment, the relative reduction in mf load was 78.1% (p<0.0001). There was no difference in the mf load 30 months post doxycycline treatment in participants who reported having or having not taken ivermectin between baseline and follow-up (Figure 1B). However, mf loads were significantly higher in the 244 participants who did not know if they took ivermectin between the two surveys or had missing data, for whom mf loads were 3.12 (95%CI: 1.73-4.52) mf/mg.

The relationship between skin infection and the number of consecutive doses missed is illustrated in Figure 2.

**Figure 2.**
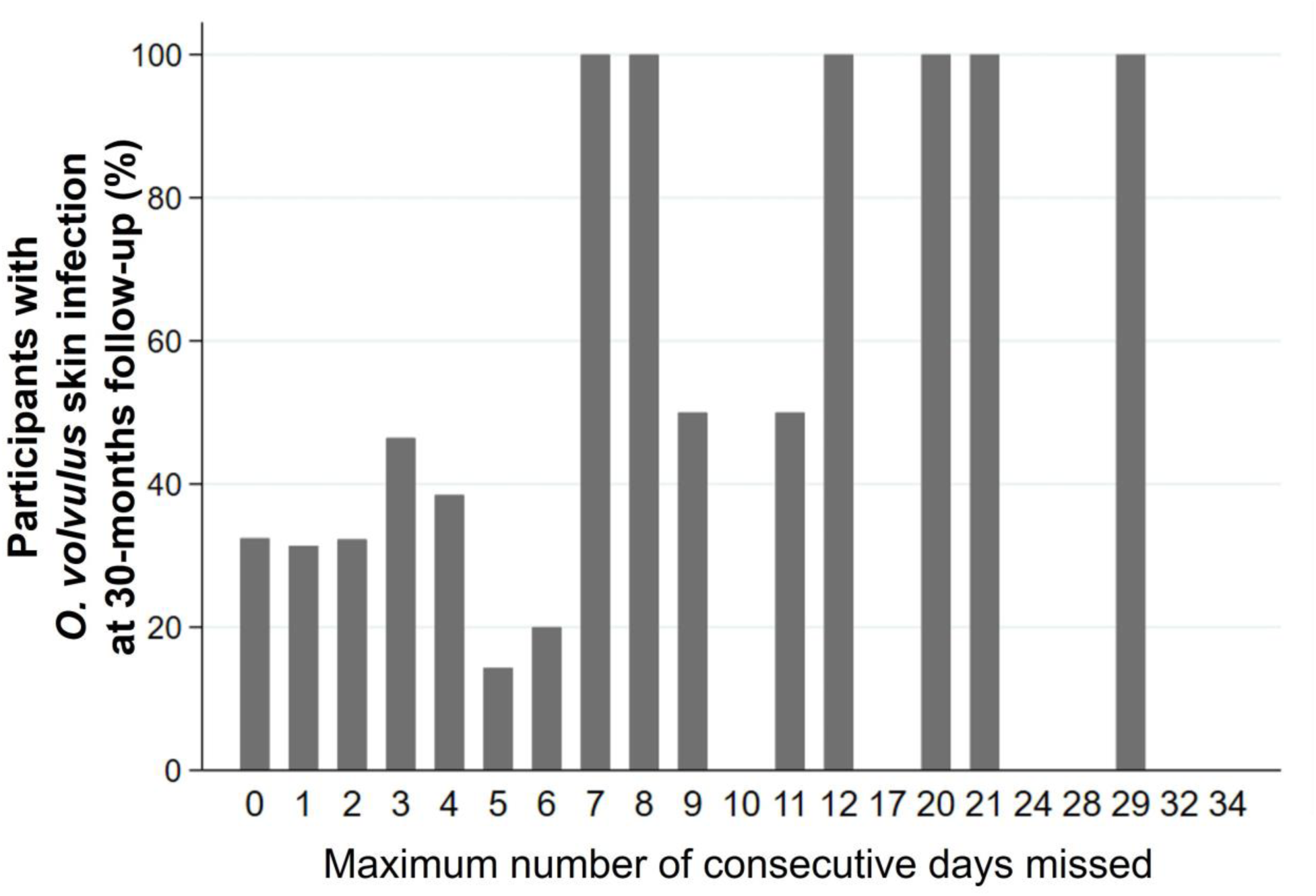
Proportion of participants with *O. volvulus* microfilaridermia 30 months post doxycycline treatment in relation to adherence to doxycycline treatment expressed as the number of consecutive doses missed in 720 participants.

The proportion of palpable nodules in all treated participants at baseline was 24.3% (95%CI: 22.8-25.9). At baseline, the number of nodules (in nodule positive participants) ranged between 1 and 7, with a mean of 1.72, a median of 1 and an interquartile range of 1. There was no difference in baseline nodule proportion (Figure 1C) or number (Figure 1E) in participants who were subsequently absent (24.5%, 95%CI: 22.8-26.3, N=2360) or present (23.8%, 95%CI: 20.8-27.0, N=719) at the 30 months follow-up (p=0.615).

The relative reduction in palpable nodule proportion 30-months post treatment was 99.0%, with only two out of 719 participants who were examined for palpable nodules having one nodule each (those two participants were nodule-free at baseline) yielding a follow-up prevalence of 0.28% (95%CI: 0.07-0.80).

### Self-reported adverse events during doxycycline treatment

Throughout the entire treatment course, adverse events were reported by 205/3,080 (6.7%) patients, and all side effects were mild. The proportion of participants reporting adverse events was similar in those present (42/720, 5.83%) or absent (162/2360, 6.86%, p=0.330) at follow up. Reported adverse events included itching (11/42, 26.2% of participants), body pain (8/42, 19.0%), weakness (7/42, 16.7%), dizziness (6/42, 14.3%), stomach pain (6/42, 14.3%), vomiting (5.42, 11.9%), swelling (3/42, 7.1%), diarrhoea (2/42, 4.8%), and headache (2/42, 4.8%).

### Factors associated with *O. volvulus* microfilaridermia at follow-up

MICE imputation method performed slightly better (FMI 46.9%) than the imputation method for monotone data (FMI 52.48%). Multiple imputation was successfully performed, with imputed values within expected range for each variable, although different from the observed values, as expected. The model estimated following imputation with the MICE algorithm is presented in Table 3. The CCA model is presented in Supplementary file 1.

**Table 3.**
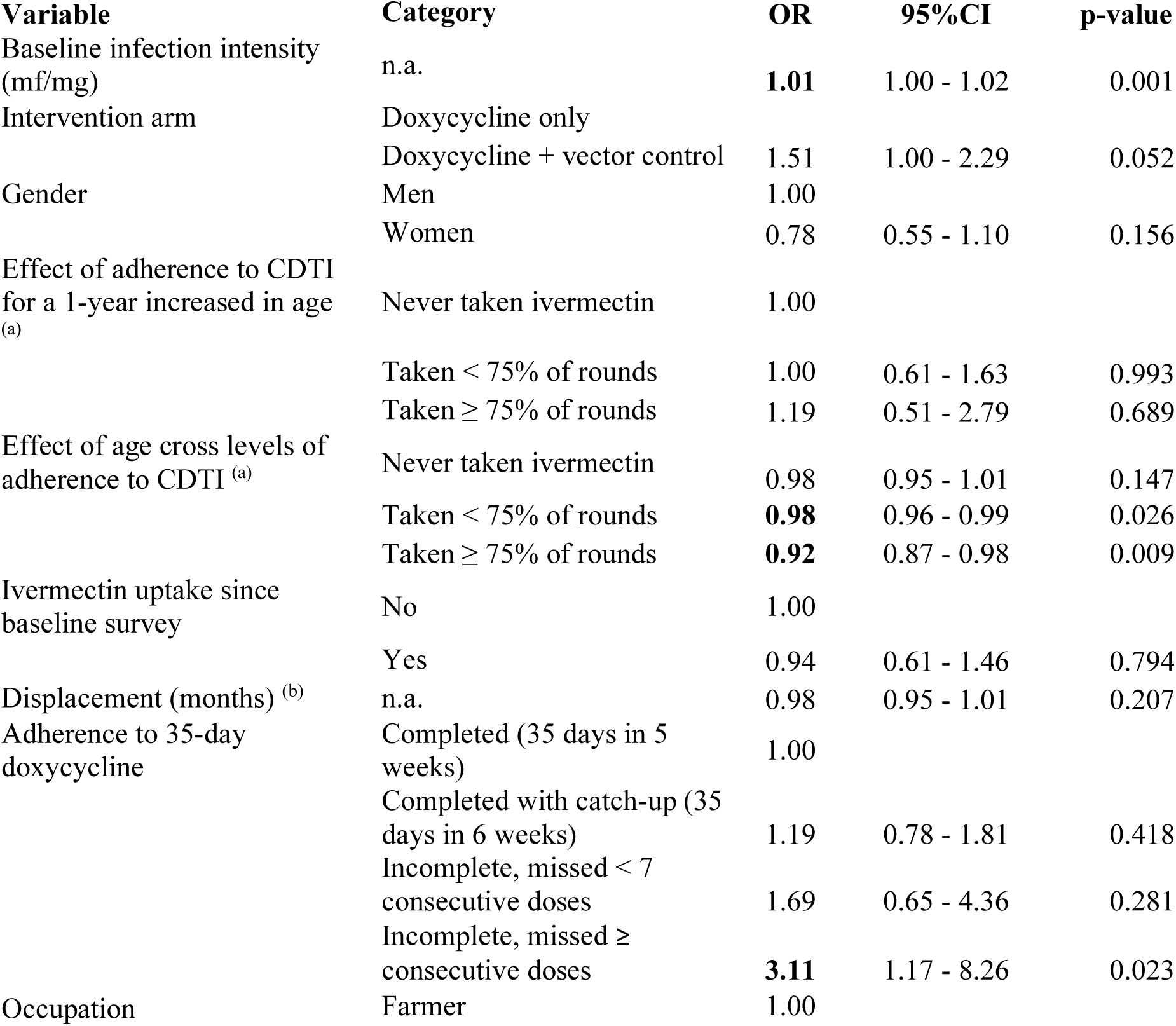
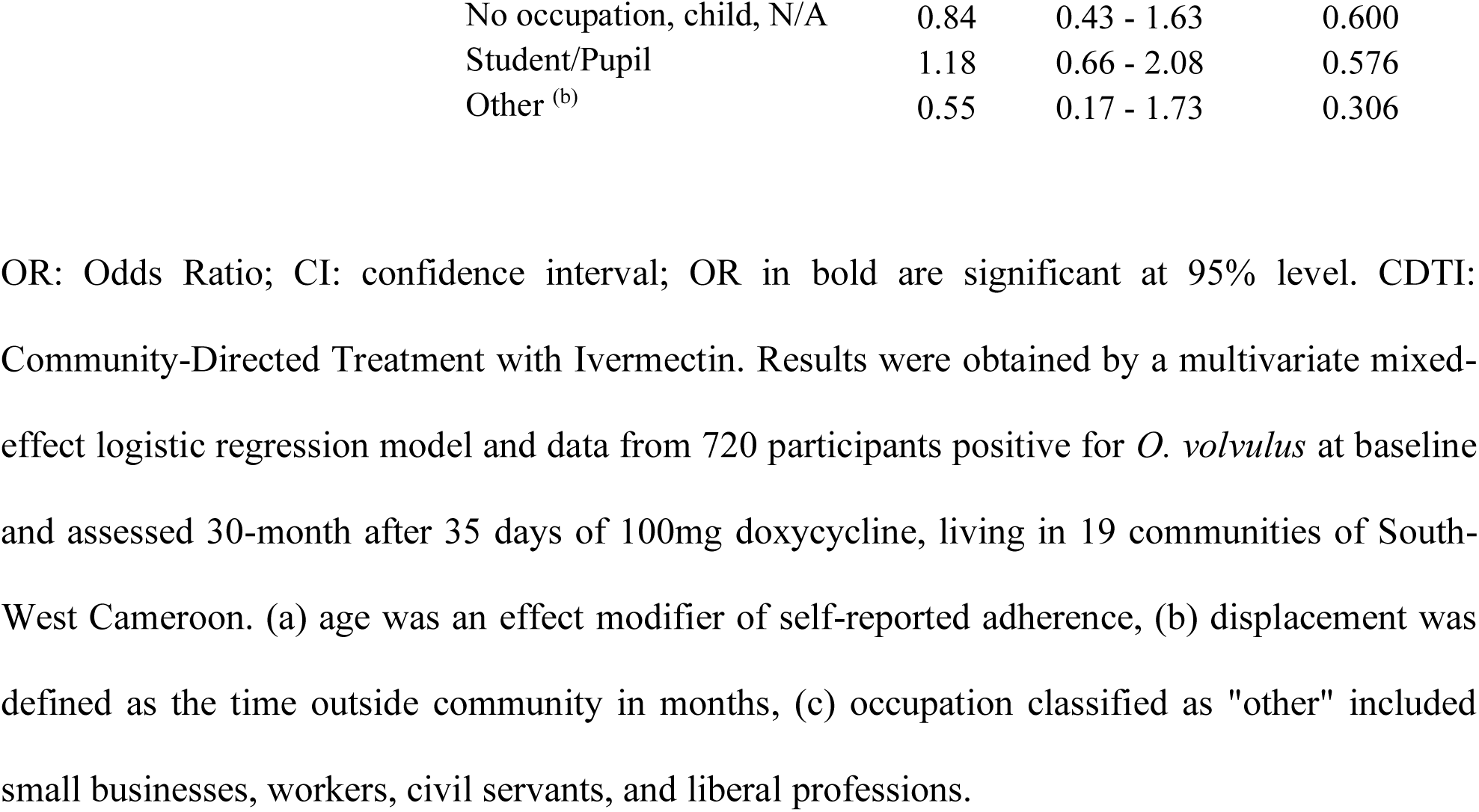
Factors associated with *O. volvulus* microfilaridermia 30-months post treatment.

The main factor associated with microfilaridermia 30-months post-treatment was having missed 7 or more consecutive daily doxycycline doses (OR 3.11, 95%CI: 1.17 – 8.26). Having missed less than 7 consecutive doses did not seem to lead to increased risk of microfilaridermia at follow-up. There was also no difference in the risk of microfilaridermia at follow-up between participants who took all 35 doses over 5 or 6 weeks. Other factors significantly positively associated with infection risk at follow-up were baseline infection intensity (OR: 1.01, 95%CI: 1.00 – 1.02), and belonging to the TTd+VC intervention arm (OR 1.51, 95%CI: 1.01 – 2.29) compared to the DOX only arm.

Ivermectin uptake since TTd treatment was not associated with risk of microfilaridermia at follow-up, nor was displacement or gender. Higher adherence to CDTI was weakly and negatively associated with microfilaridermia at follow-up (adherence to ≥ 75% of offered rounds: OR 0.93, 95%CI: 0.87-0.98; adherence to 50% to 75% of rounds: OR 0.98, 95%CI: 0.96-0.99).

## Discussion

Our analysis indicates that community-based implementation of test and treat with daily 100mg doxycycline (TTd) for 5 weeks achieved: a reduction in *Wolbachia* detection of 92% in skin *O. volvulus* mf at 6 months, a 62.8% reduction of *O. volvulus* microfilaridermia and a 99% reduction in palpable onchocercomata (adult parasite ‘nodules’) 30-months post-treatment, among 720 participants living in a high-transmission area.

The 92% depletion of *Wolbachia* in skin samples containing *O. volvulus* skin mf and/or *O. volvulus* DNA at 6-months met the empirical threshold of >90% *Wolbachia* depletion for significant macrofilaricidal activity and indicates that effective *Wolbachia* depletion efficacy was achieved in the cohort. This level of *Wolbachia* depletion is similar to levels reported in extirpated *O. volvulus* adult female worms (94.8%) at 20 months following 6-week 100mg doxycycline in a placebo controlled RCT, and to the impact on blood circulating mf four months post- doxycycline treatment in lymphatic filariasis patients ^41–44^. The ability to assess a >90% reduction in *Wolbachia* at an early interim 6-month follow-up, which is validate here as being predictive of a macrofilaricidal outcome, would be a useful early indicator of successful anti-*Wolbachia* interventions and could be used as a go/no-go decision to proceed to the extended follow-ups required for microfilaridermia and palpable nodule efficacy.

The reduction in skin mf following 5-week 100mg doxycycline is in line with findings from a prior community-based placebo controlled double-blind clinical trial in a similar population cohort in North-West Cameroon, where 6-weeks of 200mg doxycycline achieved 67% prevalence reduction at 21 months follow up ^30^. Another RCT in Ghanaian onchocerciasis patients determined an 85% reduction in skin mf prevalence after 6 weeks 200mg / day doxycycline at 20 months post doxycycline commencement ^31^. In this trial, doxycycline and placebo treated patients received ivermectin standard care 4 months after the start of treatment.

However, the 63% prevalence reduction estimated in this implementation study might be an underestimate. The follow-up survey was conducted 30 months post-treatment. While this is long enough to allow adult worm degeneration or death post-effective *Wolbachia* depletion, it increases the risk of acquiring a new infection ^31, 41, 44, 45^. With an estimated 10% annual reinfection rate post doxycycline treatment, the number of reinfections might not be negligeable in this study conducted in a high transmission area, where biting rates may reach up to 1000 infective larvae/person/month and where the two only participants found with nodules at follow-up were nodule free before doxycycline treatment ^17, 45^.

We observed a dramatic reduction in the prevalence of palpable nodules of 99% 30-months post-treatment, where all 171 patients present at follow-up who had palpable nodules at baseline were nodule-free post-treatment, and only two patients without any detected nodules at baseline were found with one nodule at follow-up presumably acquired post TTd. Prior phase II RCT trials have demonstrated loss of adult *O. volvulus* viability (curative activity) using immunohistology of aspartic protease (APR) production and ultrasound analysis of adult motility following 6-week courses of doxycycline at 21-24 months ^30, 31, 41^. With an extended 30-month timeframe, this is the first evidence of curative reabsorption of subcutaneous nodules following doxycycline treatment of onchocerciasis patients. The phenomenon of nodule reabsorption following effective tetracycline targeting of *Wolbachia* has been recorded in cattle naturally infected with *Onchocerca ochengi* after a minimal 12 month follow up period ^46, 47^.

High adherence to the 35-day treatment course, with 92.0 %, of all 3080 treated patients completing their 35-day course in 5 weeks, or with the additional, catch-up week. This was achieved thanks to extensive sensitisation, strong distribution mechanisms, i.e., observed treatment, multiple distribution points at appropriate places in each community, provision of a meal with treatment, adequate training of CDDs as well as community leaders ^48^. Previous feasibility studies of community-directed delivery of doxycycline reported similar high-levels of adherence to a six-week course of doxycycline (98%) ^39^. Community members reported relief from body itches, nodules and improved vision amongst other health benefits as reasons for adherence to treatment. Community empowerment and delivery of this alternative strategy come with additional costs compared to standard CDTi, but this must be taken into context in communities where no alternative therapeutic strategies for onchocerciasis are currently available.

Importantly, requiring an additional week to complete the 35-day course was not associated with higher risk of infection 30-months post-treatment. The odds of infection at follow-up were three-fold higher in case of 7 or more consecutive doses missed but not in case of a lower number of missed consecutive doses. Although it should be taken with caution as the number of participants in this group was small, this result suggests a fair degree of flexibility in adherence to doxycycline treatment and is in line with the minimum 4-week time frame required to achieve adequate doxycycline efficacy from modelling predictions^29^. Doxycycline was well tolerated and well accepted in those communities with low participation and high systematic non-adherence to CDTI, mostly due to the fear of side effects ^18, 37, 48, 49^.

This study has several limitations. While the low sensitivity of skin snipping is widely acknowledged, ^50–53^ there were also sensitivity issues with the qPCR for *O. volvulus* detection while assessing *Wolbachia* depletion ^40^. Most importantly, the study was originally planned as a cluster randomised controlled trial but could not be assessed as such due to civil unrest and high insecurity preventing surveying any control communities. Additionally, the follow-up survey could only be health-facility based and include participants able to travel to Kumba only.

Therefore, causality cannot be formally inferred from the available data due to the lack of controls and the impossibility to exclude selection bias due to the high loss to follow-up and recruitment at follow-up. However, we present results from a longitudinal study, and we made all possible adjustments to maximise the robustness of our findings, which we believe are representative of the study communities. Measures to mitigate bias included poststratification, to address differences in the age distribution between the baseline and the follow-up samples, adjustment for all factors associated with *O. volvulus* prevalence at baseline as well as unbalanced factors between the two surveys in the regression models, and multiple imputation to address questionnaire missing data at follow-up.

Unexpectedly, skin infection risk at follow-up, although borderline not significantly, tended to be higher in participants from communities that were targeted with ground larviciding, despite similar baseline microfilarial loads, displacement frequency and mean time, as well as self-reported ivermectin uptake after DOX treatment across the two study arms. A possible reason for this result could be the difference in the mean baseline endemicity levels between the two arms as they were represented by the community distribution in the follow-up sample^37^. Indeed, although the endemicity levels were comparable between the two arms in the baseline sample, over three quarters of participants present at follow-up were from communities with a mean *O. volvulus* baseline prevalence of 36.6% and 52.2% in the DOX and DOX *vs.* VC arms, respectively. Additionally, entomological data collected 10 weeks post larviciding indicated that Temephos suppressed biting rates at sentinel sites. However, the duration of suppression and whether this translated into reduced transmission could not be confirmed due to the impossibility to conduct entomological follow-up assessments. Therefore, no conclusion can be made about the difference in the impact of DOX vs. DOX + VC from the presented results, due to the follow-up sample imbalance in community endemicity across the two arms and the departure from the originally planned randomized control trial design and according to impact on sample size.

Doxycycline has several advantages over ivermectin for the treatment of onchocerciasis. It is curative and is completely safe in case of *L. loa* co-infection ^30, 32, 33^. Due to its benign mode of action, doxycycline does not cause inflammatory-related AEs like ivermectin, which was reflected in the present study by the high acceptability and tolerability of doxycycline ^39^. Doxycycline is a broad-spectrum antibiotic that also treats many bacterial infections and is an effective prophylactic for malaria ^54^. We acknowledge that a potential negative consequence of using this broad-spectrum antibiotic over long treatment courses and at large scale is the potential emergence of resistance to tetracyclines in other bacteria, including the gut microbiota ^55^. Whilst potential new anti-*Wolbachia* agents with shorter treatment regimens are undergoing clinical evaluation, at present, public health implementors must balance the requirements for an urgent effective curative alternative strategy for elimination of this devastating NTD where standard approaches have failed with a risk management of potential AMR impacts.

Implementing daily community-based treatment over several weeks comes with challenges and high requirements to ensure high coverage and adherence such as the presence of local health staff to reassure communities about adverse events, which is crucial in areas of current or past co-endemicity with *L. loa* and at risk of loaisis-related SAEs. It also should be noted that despite its very high efficacy, doxycycline might not suffice for an all-inclusive “endgame” scenario as it is contra-indicated in children under 9 years, or pregnant women ^34^.

This community-based intervention, together with clinical trials assessing the efficacy of doxycycline to treat onchocerciasis brings two major pieces of evidence. First, drugs targeting *Wolbachia* have high curative efficacy against *O. volvulus* ^29–31, 41–44, 56, 57^. Second, a treatment regimen longer than single-day or even multi-week treatment is feasible and efficacious following careful implementation methodologies outlined here.

The A·WOL consortium has been conducting anti-*Wolbachia* drug discovery and development since 2007, aimed at developing new anti-*Wolbachia* macrofilaricidal drugs that would require shorter treatment courses (i.e., 7 days or less), and be safe in young children and pregnant women ^34, 58–65^. Short-term treatment courses with narrow spectrum antibiotics targeting *Wolbachia* have an obvious operational advantage and would remove the selective pressure for the emergence of broad-spectrum antibiotic resistance ^34^.

The first designed anti-*Wolbachia* drug, Flubentylosin (ABBV-4083), progressed from phase I into phase II clinical studies, but lacked efficacy against onchocerciasis (clinicaltrials.gov#NCT04913610) ^58, 59^, whilst the anti-*Wolbachia* azaquinazoline, AWZ1066S, progressed into First-in-Human trials where unforeseen toxicity halted development. ^62, 63^.

Registered antibiotic repurposing of short-courses of high-dose rifampicin, rifampicin / albendazole combinations and fusidic acid, which mediate 7-14 days threshold >90% anti-*Wolbachia* efficacy in filariasis preclinical models ^59, 63^ are currently in non-regulatory proof-of-concept clinical evaluations together with ongoing hit-to-lead selection of additional fast-acting anti-*Wolbachia* compounds ^65^.

## Data Availability

All data produced in the present study are available upon reasonable request to the authors

## Acknowledgements

The authors wish to thank The Ministry of Public Health, Cameroon, South-West regional delegation of health, the district medical officers of Kumba and Konie, the chiefs of various health centres within the two health districts for their assistance and support during this field exercise. They would also like to thank the community heads and community health implementers for their support and inputs. Thanks also go to the populations of the study villages who willingly participated in this study and all those who helped in the execution of this study, particularly Mr Njato Isaac. We are thankful to Dr Caroline Jeffery for consulting on adjusting for non-participation and data imputation. This work has partially been presented at the British Society for Parasitology Spring Meeting 2019 and the 11^th^ European Congress on Tropical Medicine and International Health. We dedicate this paper to our valued colleague and dear friend, Dr Peter Enyong, who passed peacefully during its writing.

## Footnotes

### Contributors

SW, JDT, ST and MJT conceptualized the study.

SW, JDT, ST, and MJT acquired funding.

SW, JDT, LH, AJN, RAA, EDO, EK, PCN, and MJT contributed to the design of the quantitative parasitological study.

SW, MEM, LH, JDT and MJT contributed to the design of the clinical assessments.

EDO, AJN, RE, RAA, AA, TMN, EME, BN, DAN, Ste, PE, LH, FFF collected the data.

STe, AS, EEA conducted the clinical assessments.

TMN, EGF, DBN, HP and KO, analysed and interpreted the qualitative data. AF analysed and interpreted the quantitative data.

AF and EDO wrote the first draft. All authors contributed to the second draft. All the authors approved the final manuscript.

### Patient and public involvement

Patients and/or the public were not involved in the design, or conduct, or reporting, or dissemination plans of this research.

### Patient consent for publication

Not required.

### Ethics

This protocol was reviewed and approved by the Liverpool School of Tropical Medicine Research Ethics Committee (reference: 16-027), the Cameroonian National Ethics Committee for Research on Human Health (approval no. 2016/11/838/CE/CNERSH/SP), and the Division of Health Operations Research within the Cameroonian Ministry of Public Health (approval no. 631-03.17).

### Data availability

Data are available upon request. The datasets used and/or analysed during the current study are available from the corresponding author on reasonable request.

