## Supplementary File 1 for "Efficacy of test and treat with doxycycline on palpable nodules, microfilarial load and *Wolbachia* density in onchocerciasis infected persons in communities with persistent transmission in South-West Cameroon"

Factors associated with *O. volvulus* skin infection 30-months post treatment - complete case analysis (N= 451)

| **Variable** | **Category** | **OR** | **95% CI** | **p-value** |
|---|---|---|---|---|
| Infection intensity at baseline (mf/mg) | n.a. | 1.01 | 1.00 - 1.01 | 0.066 |
| Intervention arm | Doxycycline only | 1.00 | | |
| | Doxycycline + vector control | 1.45 | 0.88 - 2.4 | 0.146 |
| Gender | Men | 1.00 | | |
| | Women | 0.68 | 0.43 - 1.07 | 0.097 |
| Effect of adherence to CDTI for a 1-year increased in age [(a)] | Never taken ivermectin | 0.96 | 0.92 - 1.00 | 0.053 |
| | Taken < 75% of rounds | 0.99 | 0.54 - 1.83 | 0.980 |
| | Taken ≥ 75% of rounds | 0.57 | 0.13 - 2.49 | 0.457 |
| Effect of age in each self-reported adherence to CDTI [(a)] | Never taken ivermectin | 1.00 | | |
| | Taken < 75% of rounds | 1.03 | 0.99 - 1.07 | 0.193 |
| | Taken ≥ 75% of rounds | 1.00 | 0.93 - 1.08 | 0.935 |
| Ivermectin uptake since baseline survey | No | 1.00 | | |
| | Yes | 0.93 | 0.56 - 1.54 | 0.781 |
| Displacement (time outside community in months) | n.a. | 0.96 | 0.92 - 1.00 | 0.047 |
| Adherence to 35-day doxycycline treatment | Completed 35 days, in 5 weeks | 1.00 | | |
| | Completed 35 days with catch-up, in 6 weeks | 1.17 | 0.68 - 2.00 | 0.579 |
| | Incomplete 35 days, missed < 7 consecutive doses | 2.94 | 0.88 - 9.86 | 0.080 |
| | Incomplete 35 days, missed ≥ consecutive day | 2.75 | 0.88 - 8.56 | 0.081 |
| Occupation | Farmer | 1.00 | | |
| | No occupation, child, N/A | 0.91 | 0.40 - 2.08 | 0.83 |
| | Student/Pupil | 1.23 | 0.63 - 2.42 | 0.547 |
| | Other [(b)] | 0.61 | 0.16 - 2.27 | 0.457 |

OR: Odds Ratio; CI: confidence interval; OR in bold are significant at 95% level. CDTI: Community-Directed Treatment with Ivermectin.

Results were obtained by a multivariate mixed-effect logistic regression model and data from 451 participants with complete data, positive for *O. volvulus* at baseline and assessed 30-month after 35 days of 100mg doxycycline, living in 19 communities of Southwest Cameroon.

(a) age was an effect modifier of self-reported adherence

(b) occupation classified as "other" included small businesses, workers, civil servants and liberal professions
